# Forecasting Hospital Staff Availability During The COVID-19 Epidemic

**DOI:** 10.1101/2020.04.15.20066019

**Authors:** Chloe N. Schooling, Norbert Gyenge, Visakan Kadirkamanathan, James J.P. Alix

## Abstract

The COVID-19 pandemic poses two challenges to healthcare providers. Firstly, a high number of patients require hospital admission. Second, a high number of healthcare staff are either falling ill with the infection, or self-isolating. This poses significant problems for the staffing of busy hospital departments. We have created a simple model which allows users to stress test their rota. The model provides plots of staff availability over time using either a constant infection rate, or a changing infection rate fitted to population-based infection curves. It allows users to gauge the extent and timing of dips in staff availability. The basic constant infection rate model is available within an on-line web application (https://covid19.shef.ac.uk). As for any model, our work is imperfect. However, it allows a range of infection rates to be simulated quickly across different work patterns. We hope it will be useful to those planning staff deployment and will stimulate debate on the most effective patterns of work during the COVID-19 epidemic.

## Introduction – The Problem

Healthcare services around the world are facing their greatest challenge and this challenge has two facets. Firstly, hospitals facing unprecedented numbers of admissions. Second, they are having to simultaneously cope with reductions in staff as frontline healthcare workers are both becoming infected or self-isolating. An initial report from Wuhan, China noted that 2% of those infected were healthcare workers^1^. More recently, media outlets have reported that 5000 health staff have been infected in Italy^2^, a figure approaching 10,000 has been reported for Spain^3^. In the last few days the head of the Royal College of Physicians is quoted as saying “about one in four” of its workforce is presently off work^4^, a survey of clinicians then confirmed the high numbers of staff away from work^5^. Interestingly, in Singapore, a combination of rapid diagnosis and personal protective equipment is believed to have kept the infection rate down among health staff^6^.

Traditional medical rotas are based around experience of the unit and the number of staff required to deal safely with the workload. Those tasked with their design are not used to the rates of staff absence being reported. To try and help those involved in workforce planning we have created a model to simulate what happens to staffing levels under different infection rates. We view the model as a means to rapidly stress test staffing plans against a variety of scenarios.

### Solution - creating a model

An overview of the model is provided. The pseudocode for the model, along with the python code, is also available (supplemental files 1 and 2).

The model requires the number of staff on the rota, working pattern (e.g. a 7-day working week) and number of areas to cover. The latter might represent different areas within a department, different shifts on a ward, or different weeks on a rolling rota. The number of days off within each area can also be entered. The model will then rotate staff through different areas. Daily infection rates (%) can be entered as constant across all working areas and for both working and non-working days. Alternatively, they can be varied, for example, if one area is considered higher risk than others.

We also examine the effect of variation in daily work infection rates over time with a changing infection risk curve generated using a combination of exponential and sigmoid functions to represent the potential variations in risk over the coming months. The initial increase grows exponentially in line with the rapid growth in covid-19 patient numbers. Once the peak is reached there is a plateau for 16 days (e.g. figure 4), in keeping with reporting infection rate figures in Italy^7^ and Spain^8^. Following this a gradual decline in risk starts to occur. As the definitive rate of decline is unknown an estimation employing a mirrored sigmoid, reaching second plateau by 3 months, is used.

The expected variation in the background daily infection risk is also determined and used as the infection risk for staff on their non-working or remote-working days. This risk is calculated as the number of new cases per day divided by the size of the UK population. Assuming the UK caseload is on the cusp of its peak at the start of April, the following weeks are assumed to undertake a gradual decline in daily new cases, reaching zero by approximately 6 months. The initial rise in background risk is fitted to UK data^9^ using a least squares approach; the subsequent downturn is describing using data from Italy^10^ (21^st^ March – 7^th^ April) at 81% magnitude so that the peaks align (figure 1). Subsequent data is generated on a steady decline to reach zero by 6 months.

**Figure 1.**
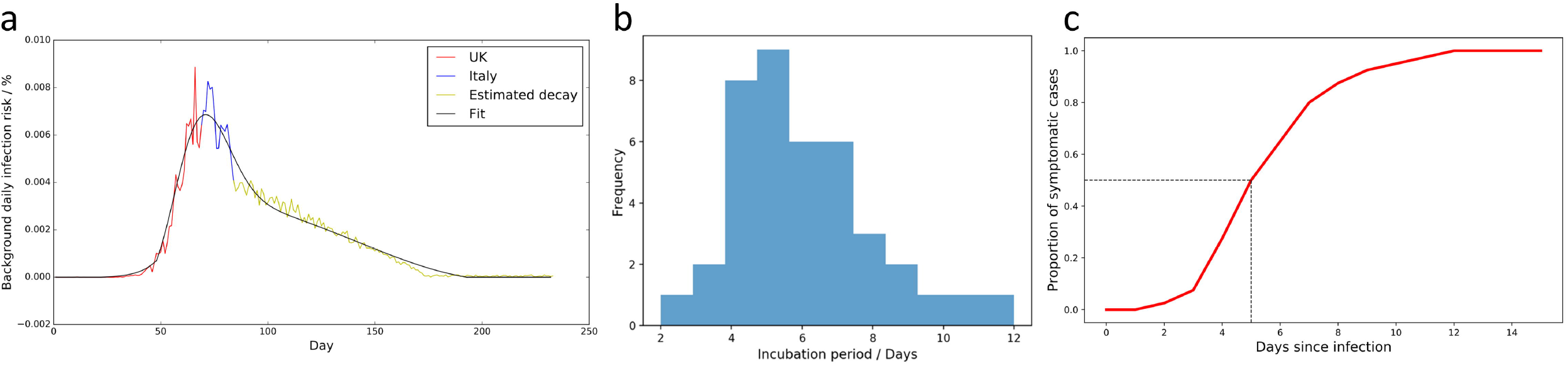
Modelling background infection risk and incubation period. a. The UK infection rate is used to produce an initial increase in background (off-work) infection risk, the downturn is fitted to emerging data from Italy. The fit is achieved using a skew normal distribution plus a third order polynomial.
b. Incubation times reported in Lauer et al^11^.
c. Distribution curve for the incubation period fitted to the Lauer et al data.

Self-isolation may also be incorporated into the model and this is done using a risk of infection and self-isolation in a 2:1 ratio, figure based on the reported ratio among physicians in the UK^5^. Any staff member who self-isolates will be unavailable to work for 14 days.

There remains a risk of infection while self-isolating, which we deem higher than the general background risk and is arbitrarily set at a daily rate of 0.11% (which accumulates to approximately 20% being infected during the 2-week isolation period).

As staff progress through the rota the model uses random number generation in line with the infection risk each day, in order to determine if, and when, each staff member becomes infected. Within the model the incubation time between exposure and infection is set to a median of 5 days with a range of incubation times arrived through fitting to data from Lauer et al^11^ (figure 1). After symptom onset, 80% of staff have an illness that allows them to gradually return to work after 14 days. 97% of the workforce is back after 6 weeks post-symptom onset and they are gradually re-introduced. 3% do not return to work within the time period of the analysis. The model is run as a Monte Carlo simulation through repeated sampling and the resulting mean and 95% confidence intervals for staff levels are plotted.

In addition, it is possible to specify a critical area in which staff levels must not drop below a specific number. Staff are moved from other areas to support the critical area and outputs concerning the number of consecutive days worked, percentage of days off work within the rota and the number of days redistributed to the critical area are reported. This redistribution is run first with no specific criteria and then with an assignment policy in order to spread out the intensity of work. Staff are selected to move based on how many times they have previously been redistributed to the critical area, with a secondary condition on the number of consecutive days they have worked in a row (up to a maximum of 8).

A simple web application which implements the constant infection rate algorithm is available at https://covid19.sheffield.ac.uk.

## Results

We present results for two departments. One uses a proposed shift system for an acute medical team, based on a proposed shift system for the NHS Nightingale Hospital, London. The other is based on an approach explored locally for the delivery of a highly specialised tertiary service. We first present relatively simple scenarios that can be run within our presently available online web application. We then describe a more complex setting.

### Constant daily risk: acute medicine

The shift system is based around a 7-day working pattern and utilises a total of 20 staff. While in reality these would be split into multiple teams working different shifts on different days, the pattern can be simplified to one group (table 1).

**Table 1.** Acute medical team working pattern.

| Shift 1 | Shift 2 | Shift 3 |
| --- | --- | --- |
| 2 long days | 2 nights | 3 days off |

We first consider the at work and off-work daily infection risks, utilising reported figures for NHS staff infections and overall population risk. Firstly, on day 52 of the UK covid-19 crisis the Secretary of State for Health and Social Care quoted a figure of 6% for the proportion of NHS staff off due to covid-19 infection^12^. The off-work infection risk represents the overall population risk of infection. There are well documented limitations to the ongoing approach to testing and so we considered two countries, the UK^13^ and Germany^14^. For the period 31st March - 6th April both countries, interestingly, returned an average daily infection rate of 0.0063%.

While there are different constant daily infection rates that can produce a staff absence of 6% on day 52 of the model, we present two contrasting approaches: a low-grade approach with a daily infection risk of 0.45% and a more pronounced 14.2% daily risk. In the low-grade model a fairly constant absence of staff is seen from around day 25 with an average of 18.5 staff available (95% confidence interval 15.2-20; figure 2a). Under the more severe daily risk of 14.5%, the effect is observed much earlier and in a more severe fashion; the peak is observed at day 18, with an average of 3.5 staff available (figure 2b). However, the duration in which large numbers of staff are absent is much shorter; a 50% reduction is staff is observed between day 10 and 26, with a return to steady state by day 60.

**Figure 2.**
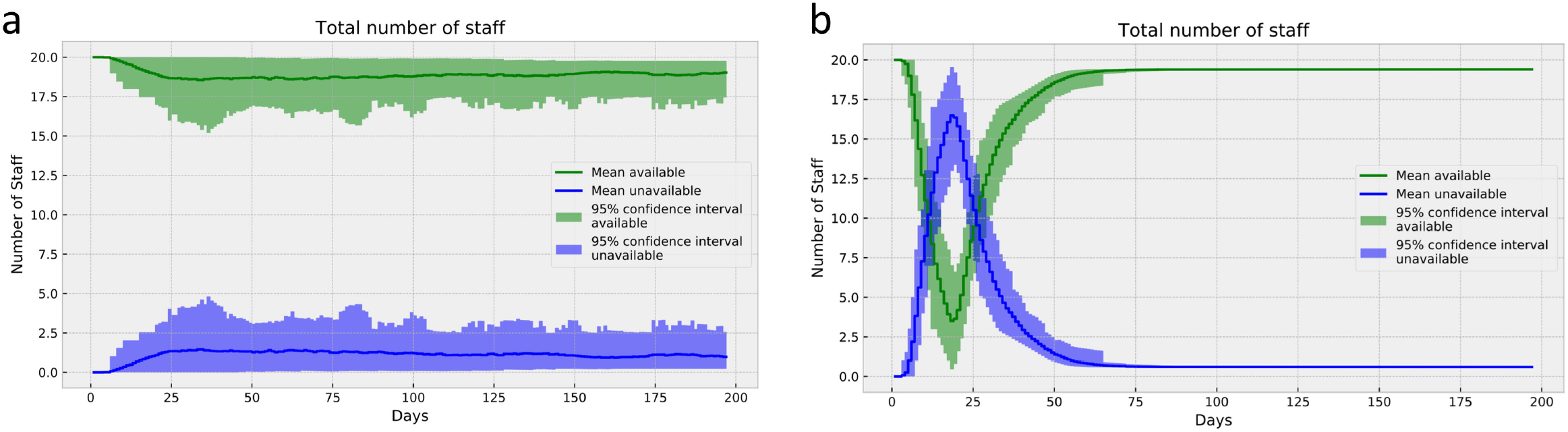
Staff levels for the acute medicine team with different at work infection rates. Both models result in 6% of staff being absent on day 52 but do some through different at work infection risks. a. A constant low-level infection risk (0.45%/day) produces a long plateau of staff absence.
b. A higher rate (14.5%/day) results in a more severe but shorter dip in staffing levels. The 6% absence rate on day 52 is now largely achieved by staff returning to work after infection.

### Constant daily risk: tertiary specialised service

Here we simulate having 36 doctors available with different services to deliver: acute ward-based work, urgent face-to-face clinic appointments and telephone clinics (table 2). The shift pattern is based on the 7-day week, however, in areas 2 and 3 staff work 5 days only (e.g. Monday-Friday). The inpatients at night are covered by a “hospital at night service”, with remote specialist advice available.

**Table 2.** Tabulation of the tertiary neurology service.

| Area 1 (Mon-Sun) | Area 2 (Mon-Fri) | Area 3 (Mon-Fri) |
| --- | --- | --- |
| Acute ward work<br>*2 teams:*<br>*4 days on/3 days off*<br>*4 days off/3 days on* | Face-to-face clinics | Telephone clinics |
| Daily work risk:<br>6.2% | Daily work risk:<br>0.11% | Daily work risk:<br>0.0063% |
| Daily off work risk: 0.0063% |  |  |

Here, we demonstrate a different approach to the distribution of infection risk. A report on the European Academy of Neurology website (www.ean.org) described the experience of the neurology unit in Brescia, Italy, in which 8 out of 18 neurologists (44%) were infected in 32 days^15^. Using the risks shown in table one results in this number of infections by day 32.

Using this model, we derive staffing distributions with a 3-day nadir of 15.7 staff available between days 19 and 22 (figure 3, a1-4). As the acute service can be considered critical, we then protect the staff levels in this area which leads to reductions in the staff available to other areas (figure 3, b1-4).

**Figure 3.**
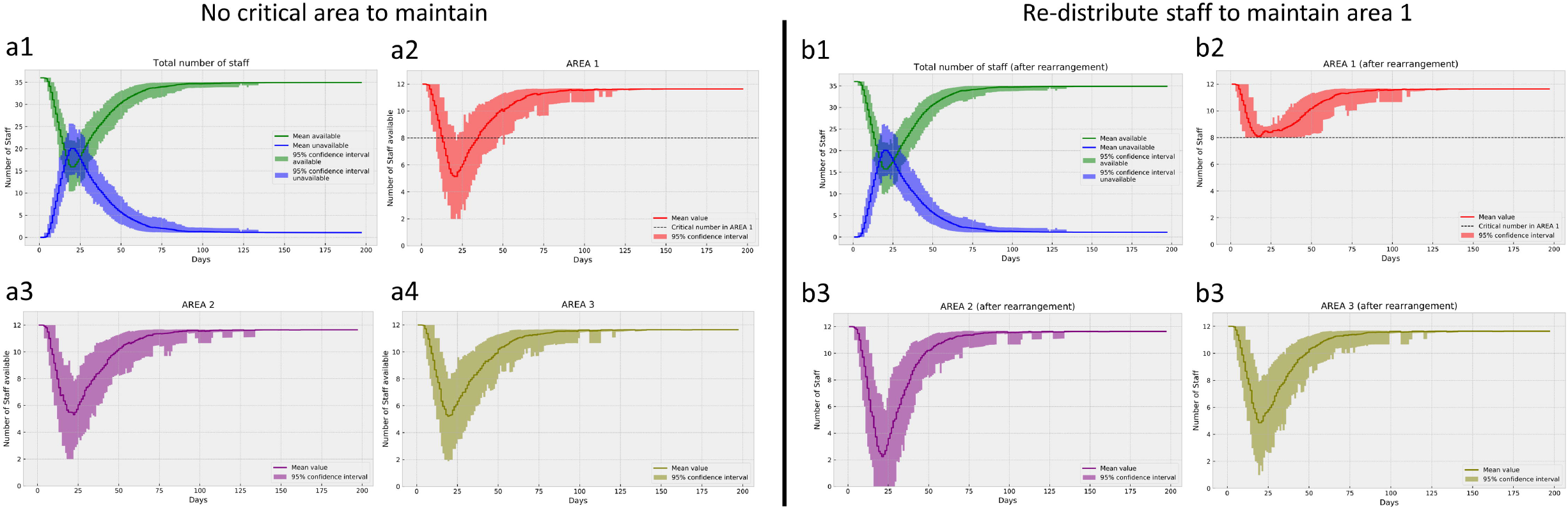
Modelling multiple areas with differing infection risks. a. 1-4: Different daily risks are attributed to the three areas but as staff rotate through each area the higher risk work drives staff absence across areas.
b. 1-4: A redistribution of staff occurs in order to maintain staff levels at n=8 in area one (acute ward based work).

### Modelling a changing infection risk

We now consider a changing infection risk for both at work and off-work time periods, using two quoted figures of NHS staff illness at day 52 of the UK crisis, 6%^12^ and 25%^4^ (see methods).

For the acute medical team, under the assumptions of the 6% staff illness model, staffing levels reach their lowest point between days 54 – 60, with an average of 17.7 staff available (95% confidence interval 13.9 – 20; figure 4). Adjusting for 25% of staff being infected on day 52 (and hence off work) results in a more severe drop in staff (nadir average of 13.4 available on day 56, 95% confidence interval 16.6 – 8.9). However, the peak is shorter in duration lasting 3 days (day 54 – 57).

**Figure 4.**
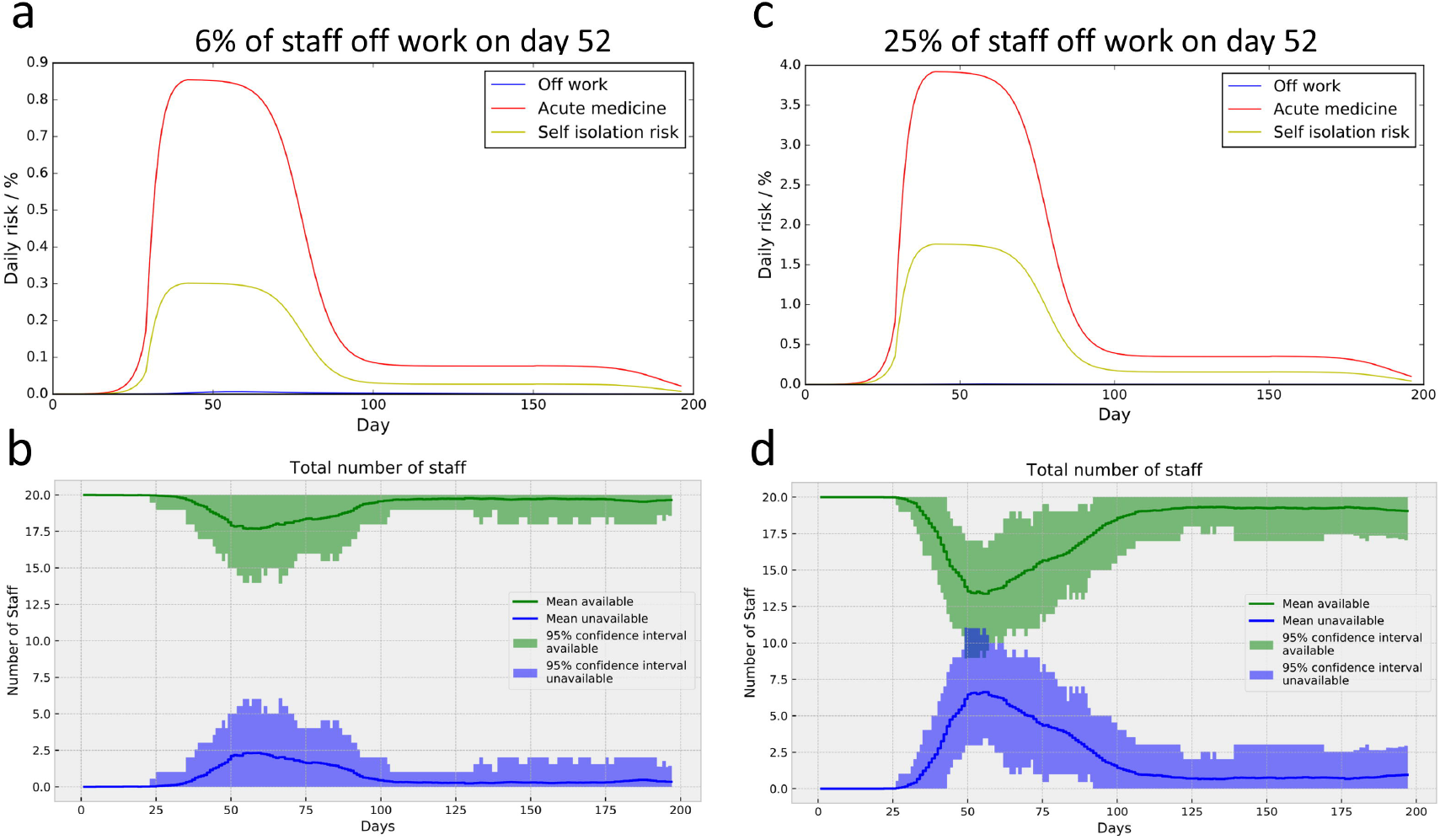
Modelling changing infection risk for acute medicine team. a. Daily risk for different work areas over time arriving at 6% of stay being off work due to infection on day 52 of the model.
b. A relatively low-level loss of staff is seen.
c. Daily risk for different work areas over time arriving at 25% of stay being off work due to infection on day 52 of the model.
d. A more severe loss of staff is seen over a similar time period to that observed in (b).

For the tertiary department trying to maintain both inpatient and outpatient work, a 6% staff infection figure at day 52 results in lowest staffing levels on days 51-56 (average of 32.5 available, 95% confidence interval 29 – 36; figure 5). Increasing to 25% of staff off on day 52 results in a greater magnitude of staff loss but (an average of 21.8 staff available, confidence interval 17 - 28, day 55).

**Figure 5.**
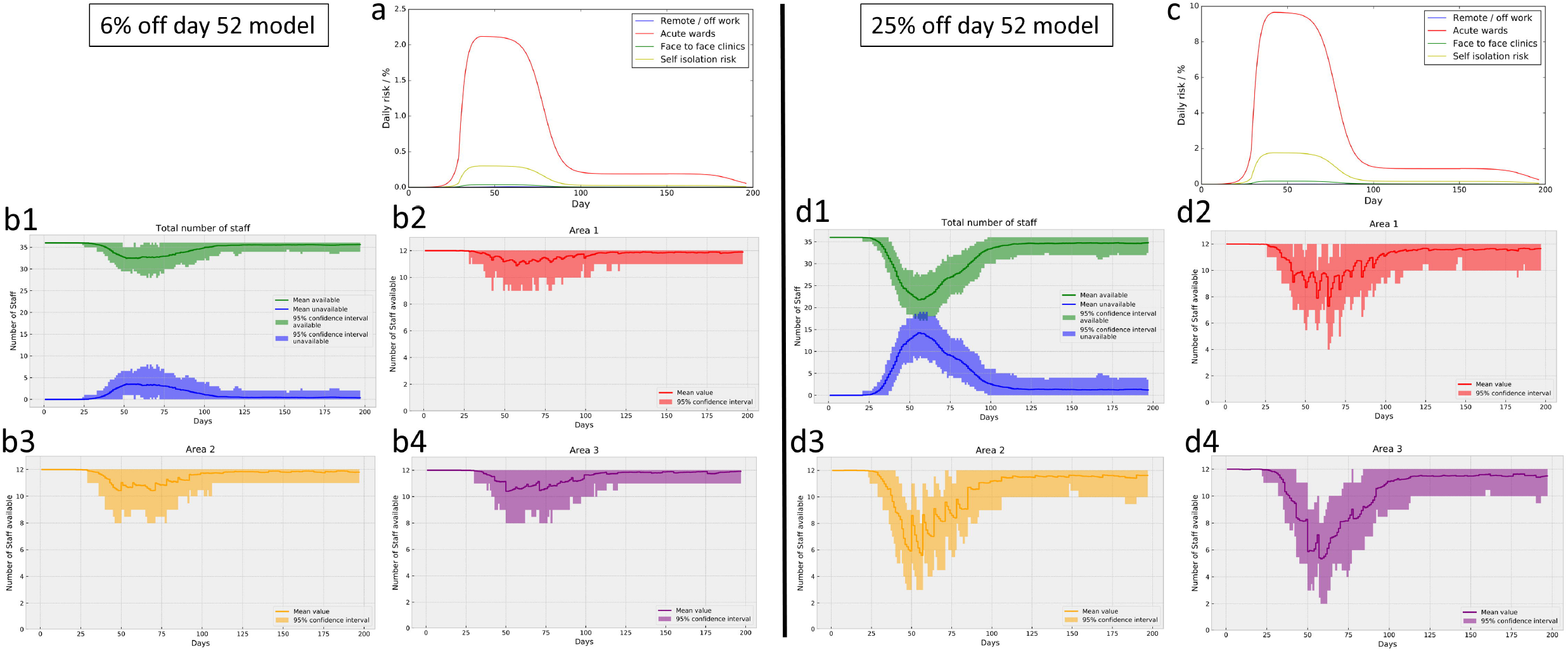
Modelling changing infection risk for tertiary medical team. a. Daily risk for different work areas over time arriving at 6% of stay being off work due to infection on day 52 of the model.
b. 1-4: A relatively low level loss of staff is seen across all areas of work.
c. Daily risk for different work areas over time arriving at 25% of stay being off work due to infection on day 52 of the model.
d. 1-4: A more severe loss of staff is seen.

Staffing levels in the acute work areas could be supported through redistribution of staff (figure 6). When supporting acute ward-based work as a critical area with a 25% staff absence on day 52, we examined the effects of setting criteria for the selection of staff. Staff levels were maintained both with and without conditions, but the latter resulted in reductions in the number of consecutive days worked and maximum number of times each individual was redistributed (figure 7).

**Figure 6.**
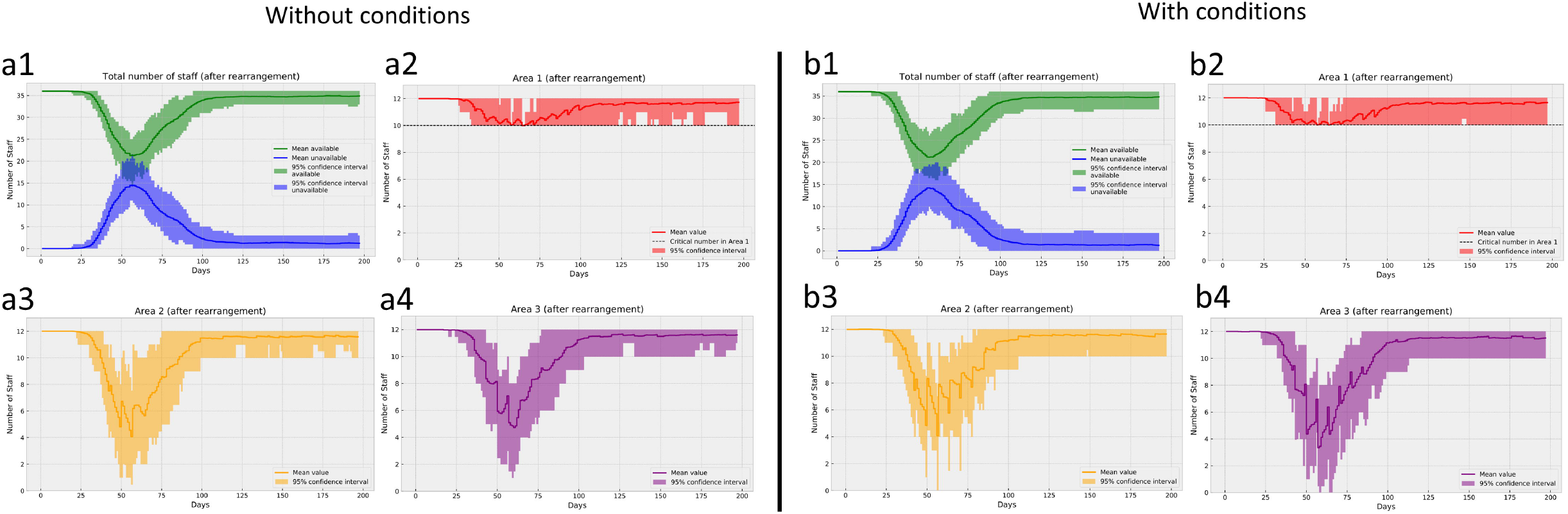
Preserving acute care team within a tertiary medicine model. With 25% of all staff unavailable on day 52 the acute care area (area 1) can maintain and high staff levels through redistribution of staff from other (non-acute) areas without (a1-4) and with (b1-4) conditions governing the re-distribution.

**Figure 7.**
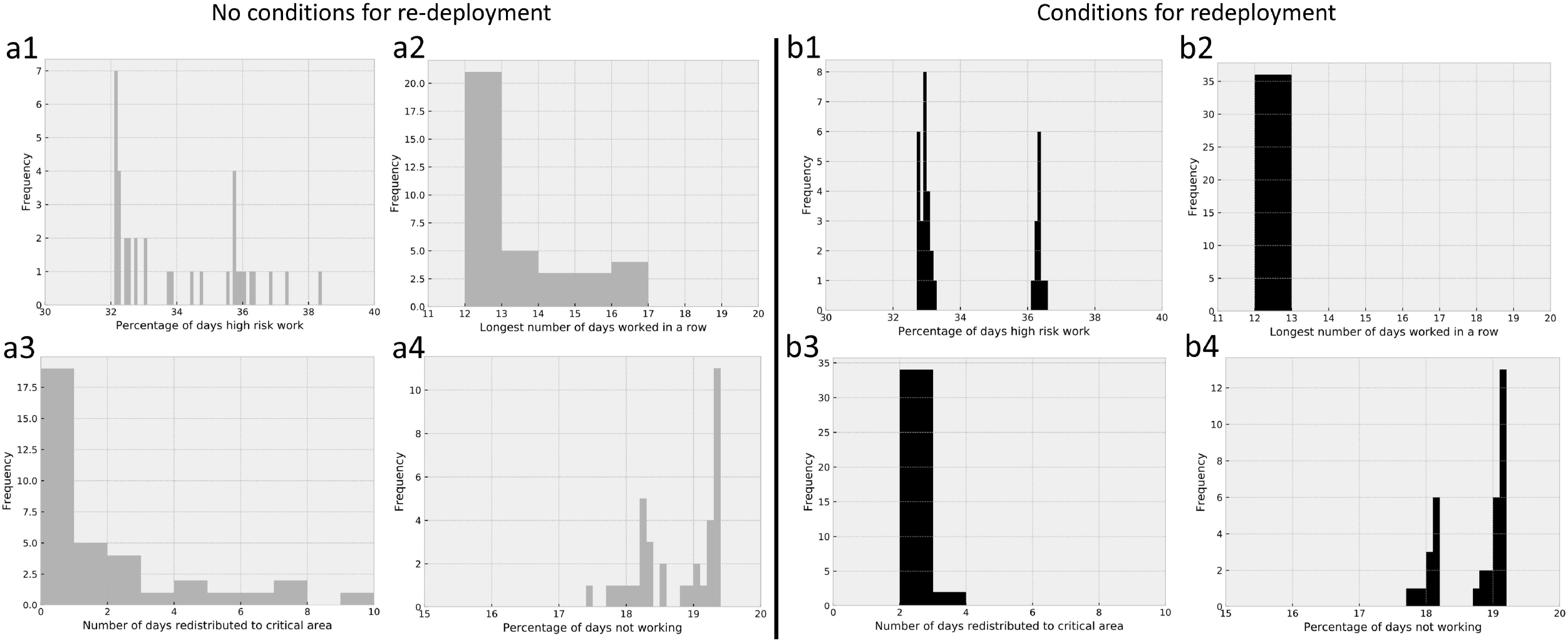
Working conditions without and with conditions for redistribution of staff. a. 1-4. Frequency histograms for work done by staff as some are redistributed to support the acute care group in figure 6 without any conditions governing the pattern of work allowed. Half of all staff (n=18) are not redistributed at any time (a3).
b. 1-4. Results following use of simple rules for the redistribution of staff demonstrated a reduction in important working conditions. Now all staff are redistributed at some point, but the number of days spent in the new group are much reduced (b3).

## Discussion

We present a simple model and provide examples of how it can be used to stress test medical staff rotas within a variety of different scenarios. The model produces information on the extent and timing of the loss of staff. If the infection risk is immediate and constant, our data suggest staffing levels dip from around 20 days after the onset of staff exposure. If the infection risk increases in line with the present rate of infection seen in populations, then we observe a staffing nadir around day 55 of the local epidemic.

In ideal circumstances, the risk to health professionals would be zero and they could focus solely on the care of their patients. Unfortunately, this has not been observed and frontline staff are withdrawing from clinical duties due to sickness. Our approach offers some insight into the potential effect of staff illness on staff levels over time. The model allows different scenarios to be modelled quickly and, while imperfect, may enable healthcare leaders to consider different circumstances and plan mitigation strategies.

We simulate both constant and changing infection rates and we appreciate that there are many unknowns in this regard. So far, different countries have experienced quite different levels of infection, both at a population level, and also for healthcare staff. The reasons for this are not definitively known but may relate to the approach to contact tracing, self-isolation, PPE, among others. Similarly, while we have used the best available evidence we could find, we cannot be sure how the infection rate changes over time among healthcare staff and how different interventions interact. For example, the risk of infection from working in a ward with few covid-19 positive patients may be different to the risk encountered by the same staff working on the same ward when/if all patients are covid-19 positive. Intuitively, one might think that the latter poses a higher risk, however, a greater use of PPE, or increased staff experience (for example), could result in the opposite effect.

Infection rates may also vary between different types of hospital environment e.g. A+E/ITU/medical ward, again this is unknown. Similarly, infection risk outside of the hospital may also be different across different countries, as well as within different regions of an individual country, perhaps relating to factors such as population demographics, density and the availability of medical facilities, although there are many potential factors. By using available statistics on the background population risk (R_B_), together with the time period of encountering covid-19 positive patients (T_tot_), the number of days off work within the time period (T_off_) and the number of staff who have been infected (N_i_), one can estimate an “at work” infection rate (R_W_). Mathematically this may be expressed as:

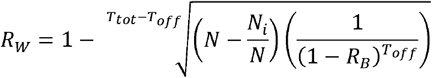

In our scenarios we introduce the notion of a critical area, one for which staffing levels must not drop below a certain level. In practical terms, this may occur within a department, as demonstrated in our tertiary specialised medicine example, or at the level of a whole department (e.g. an intensive care unit). Resource allocation is well studied and there are many different approaches available. As the redistribution of staff may incur a “cost”, that is, significant changes in work patterns such as duration and intensity, we introduced a simple assignment policy to reduce such effects. Such systems could be implemented with the use of feedback loops to optimise staff allocations (e.g.^16^).

Lastly, we have not discriminated between different levels of doctor within our existing model, although clearly the option to develop this exists. In the UK, guidance has been issued on how to group staff at different grades^17^. Often rotas are done for each grade separately and so our present analysis focuses on a single staff group.

In summary, we describe a simple approach to rapidly examine the potential effects of different staff infection rates on healthcare rotas in the covid-19 pandemic. We hope our results will stimulate discussion on this important area.

## Data Availability

Pseudocode and phython script are provided.

## Acknowledgements

We thank Dr Daniel Blackburn and Dr Matthew Roycroft for their discussion on rota patterns.

